# The effect of climate change on Yellow Fever disease burden in Africa

**DOI:** 10.1101/2020.02.25.20026369

**Authors:** Katy Gaythorpe, Arran Hamlet, Laurence Cibrelus, Tini Garske, Neil Ferguson

## Abstract

Yellow Fever (YF) is an arbovirus capable of causing haemorrhagic fever which is endemic in tropical regions of Africa and South America. In recent years, it has resurged – leading to large outbreaks and expanding its endemic zone, the causes of which are unknown. In Africa, the disease is currently considered endemic in 34 countries where it is estimated to cause 78,000 deaths a year. As the mosquito vectors of YF sensitive to environmental conditions, climate change may have substantial effects on the transmission of YF. Here we present the first analysis of the potential impact of climate change on YF transmission and disease burden. We extend an existing model of YF transmission in Africa to account for rainfall and a temperature suitability index. From this, we project transmission intensity across the African endemic region in the context of four climate change scenarios (representative concentration pathways (RCPs) 2.6, 4.5, 6.0 and 8.5). We use these transmission projections to assess the change from current to future disease burden in 2050 and 2070 for each emission scenario. We find that disease burden changes heterogeneously with temperature and rainfall across the region. In RCP 2.6, we find a 93.0% [95% CI 92.7, 93.2%] chance that deaths will increase in 2050. We find that the annual expected number of deaths may increase by between 10.8% [95% CrI -2.4, 37.9%] for RCP 2.6 and 24.9% [95% CrI -2.2, 88.3%] for RCP 8.5 in 2050, with the most notable changes occurring in East and Central Africa. Changes in temperature and rainfall will affect the transmission dynamics of YF. Such a change in epidemiology will complicate future control efforts. As such, we may need to consider the effect of changing climactic variables on future intervention strategies.

## 1 Introduction

Yellow Fever (YF) is a zoonotic arbovirus endemic in tropical regions of Africa and Latin America. It is responsible for approximately 78,000 deaths per year although under reporting is high and since YF has a non-specific symptom set, misdiagnosis is an issue (Garske et al. 2014). YF has three transmission ‘cycles’ in Africa: urban, zoonotic and intermediate. The urban cycle, mediated by *Aedes Aegypti* mosquitoes, is responsible for explosive outbreaks such as the one seen in Angola in 2016 (Ingelbeen et al. 2018; Wilder-Smith and Monath 2017). While the urban cycle can rapidly amplify transmission, the majority of YF infections are thought to occur as a result of zoonotic spillover from the sylvatic reservoir in non-human primates (NHP). This zoonotic cycle is mediated by a variety of mosquito vectors including *Aedes Africanus* and, as the NHP hosts are mostly unaffected by the disease in Africa, the force of infection due to spillover is fairly constant, unless land use changes (Monath and Vasconcelos 2015). The intermediate cycle is sometimes called the savannah cycle and is mediated by mosquitoes such as *Ae. luteocephalus*, who feed opportunistically on humans and NHP (Barrett and Higgs 2007).

The Intergovernmental Panel on Climate Change (IPCC) states that global mean temperatures are likely to rise by 1.5°C, compared with pre-industrial levels, by between 2030 and 2052 if current trends continue (Masson-Delmotte et al. 2018). Increases are projected not only in mean temperature but in the extremes of temperature, extremes of precipitation and the probability of drought (Kharin et al. 2013; Dunning, Black, and Allan 2018).

With multiple mosquito vectors and a zoonotic cycle depending on NHP hosts, the impact of climate change on YF is likely to be complex. Focusing on the main urban vector, *Ae. Aegypti*, there is strong evidence that projected climate change will alter its global distribution and thus, the risk of diseases it carries (Ryan 2019; World Health Organisation 2018). Climate change has been predicted to increase the regions at risk from dengue and Zika transmission, though seasonal variation in temperature may mitigate the likelihood of outbreaks ion areas at the edges of the endemic zone (Mordecai et al. 2017; Huber et al. 2018).

Long-term projections of the future disease burden of YF are needed to inform vaccination planning (VIMC 2019). Furthermore, differences due to climate change may increase the risk of epidemics, a key consideration for the Eliminate YF Epidemics (EYE) strategy (World Health Organization 2017).

In this manuscript we extend an existing model of YF occurrence and disease burden to incorporate a nonlinear temperature suitability metric. This measure of temperature suitability depends on the thermal responses of the vector, *Ae. Aegypti*, and virus which we estimate specifically for YF. We combine this with YF occurrence data in a Bayesian hierarchical model in order to account for uncertainty at each stage of the modelling process. This, along with established estimates of transmission intensity informed by serological survey data, allow us to predict current and future transmission intensity. Finally we use ensemble climate model predictions of future temperature and precipitation to project transmission and thus, burden in 2050 and 2070. Our results are the first examination of YF burden under the effect of climate change.

## 2 Materials and methods

### 2.1 Datasets

We use a number of data sets to inform both the generalised linear model (GLM) of YF occurrence and the temperature suitability model. Additionally, we rely on estimates of transmission intensity informed by serological studies which are detailed in Gaythorpe et al. (2019) and described below.

#### 2.1.1 YF occurrence

Details of YF outbreaks occurring from 1984 to present day were collated into a database of occurrence. These data were collected from the World Health Organisation (WHO) weekly epidemiological record (WER), disease outbreak news (DON), published literature and internal WHO reports (World Health Organization 2009; World Health Organization, n.d.). Additionally, reports of suspected YF cases were collected in the WHO African Regional Office YF surveillance database (YFSD); this included data from 21 countries in West and Central Africa. The database was based on the broad case definition of fever and jaundice leading to a large proportion of cases attributed to non-YF causes. However, the incidence of suspected cases can be used as a measure of surveillance effort and is included as a covariate in the generalised linear model.

#### 2.1.2 YF serological status

Surveys of seroprevalence were conducted in Central and East Africa. We use these to asses transmission intensity in specific regions of the African endemic zone. The current study includes surveys from published sources (Diallo et al. 2010; Kuniholm et al. 2006; Merlin et al. 1986; Omilabu et al. 1990; Tsai et al. 1987; Werner, Huber, and Fresenius 1984) and unpublished surveys from East African countries conducted between 2012 and 2015 as part of the YF risk assessment process (Tsegaye et al. 2018). The surveys were included only if they represent the population at steady state, as such outbreak investigations were omitted (Garske et al. 2014). Additionally, in the majority of surveys, vaccinated individuals were not included; however, in South Cameroon, vaccination status is unclear an so we fit an additional vaccine factor for this survey. Summary details of the seroprevalence studies are included in the supplementary material.

#### 2.1.3 Past vaccination coverage and demography

Vaccination coverage is estimated using data on historic large-scale mass vaccination activities (Durieux 1956; Moreau et al. 1999), routine infant immunization reported by the WHO and UNICEF (World Health Organization/ UNICEF 2015), outbreak response campaigns detailed in the WHO WER and DON and recent preventive mass-vaccination campaigns carried out as part of the yellow fever initiative (World Health Organization 2009; World Health Organization, n.d.). The coverage is estimated with the methodology of Garske et al. and Hamlet et al. and is visualised in the polici shiny application (Garske et al. 2014; A Hamlet, Jean, and Garske 2018). The application provides vaccination coverage estimates at province level for 34 endemic countries in Africa which can be downloaded for years between 1940 and 2050.

Demography is obtained from the UN World Population Prospect (UN WPP) (United Nations DoE 2017). We dis-aggregate this to province level by combining it with estimates of spatial population distributions from LandScan 2014 (Dobson et al. 2000). This allows us to estimate population sizes at province level for each year of interest assuming that the age structure is relatively similar across all provinces in each country.

#### 2.1.4 Environmental and climate projections

We use three main environmental covariates within the generalised linear model of YF occurrence: mean annual rainfall, average temperature and temperature range, shown in figure 1. These are gridded data at various resolutions, ranging from approximately 1km to 10km, which we average at the first administrative unit level (NASA 2001; Xie and Arkin 1996; Hijmans et al. 2004).

**Figure 1:**
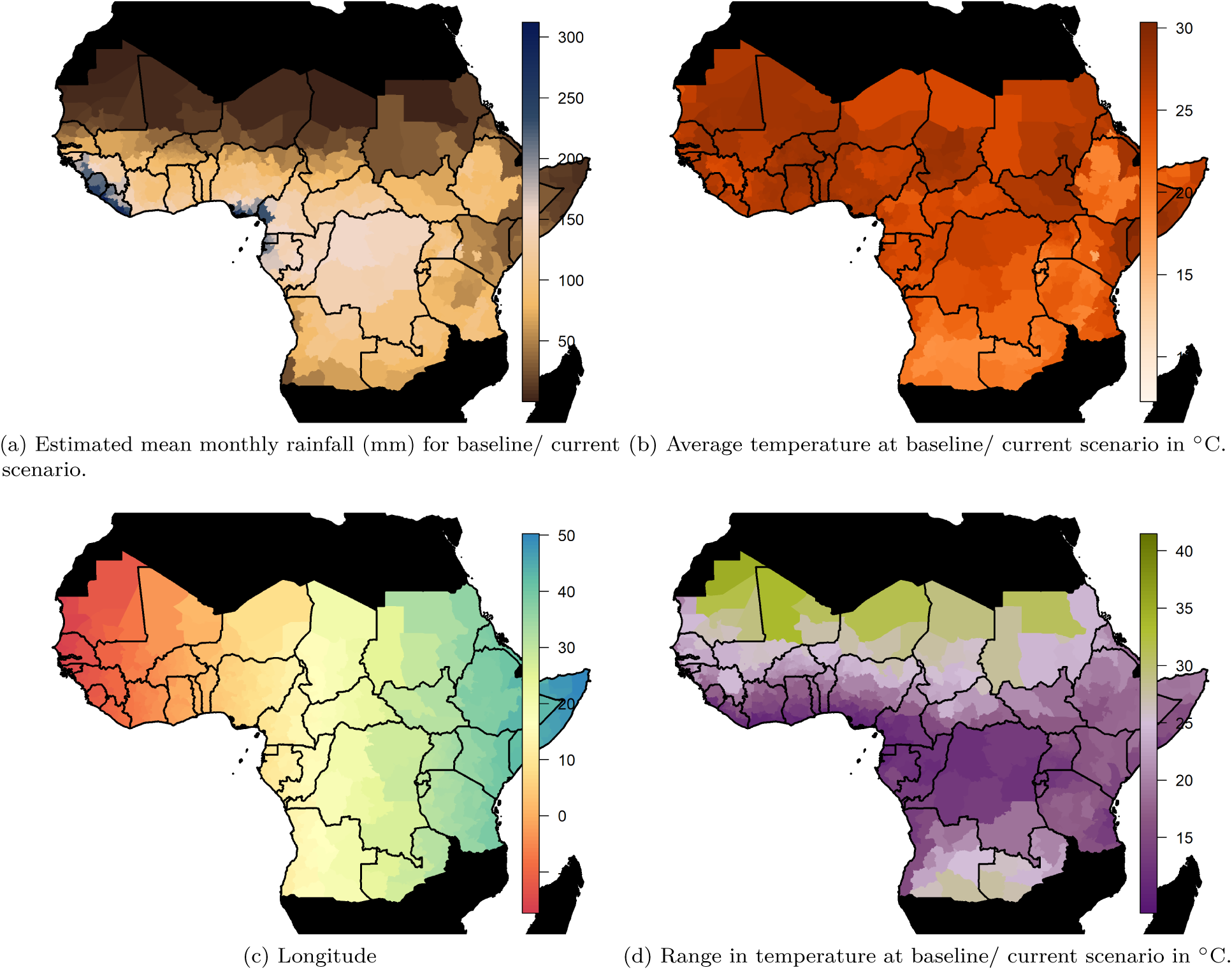
Spatial data inputs for generalised linear model. Countries shown in black are not considered endemic for YF.

Projected temperature and rainfall changes under climate change scenarios were obtained from worldclim version 1.4 (Hijmans et al. 2005; Fick and Hijmans 2017). These data provided the 5th Intergovernmental panel on climate change (IPPC5) climate projections for four Representative Concentration Pathways (RCPs): 2.6, 4.5, 6.0 and 8.5 (Van Vuuren et al. 2011). The different RCPs indicate different possible emission scenarios and represent the resulting radiative forcing in 2100, measured in W/m_2_ or watts per square metre. Each scenario is assumed to peak at a different times, with emissions peaking between 2010 and 2020 for RCP 2.6, but rising throughout the century for RCP 8.5. Projections of the mean global temperature rise by 2046 - 2065 are 1 or 2 °C for RCPs 2.6 or 8.5 respectively, compared to pre-industrial levels of the 1880s. By the end of the century, these projections suggest a rise of 1 [0.3 to 1.7] or 3.9 [2.6 to 4.8]°C for RCPs 2.6 or 8.5 (Stocker et al. 2013; Rogelj, Meinshausen, and Knutti 2012). Current warming is estimated to be 0.85 °C since pre-industrial levels (Stocker et al. 2013). Based on current commitments through aspects such as the Paris agreement, scenarios where temperatures are expected to rise by more than 3 °C have been suggested to be most likely (Sanford et al. 2014). As such, a recent study omitted the RCP 2.6 scenario as it is unlikely now to occur (Mora et al. 2013; Vliet, Elzen, and Vuuren 2009).

Projected mean rainfall, maximum temperature and minimum temperature are available for each RCP scenario in years 2050 and 2070. We take the midpoint and range of the temperature as inputs for the model of YF occurrence, where the midpoint temperature is used to calculate the temperature suitability index. A comparison of the temperature mean and midpoint for the baseline/ current scenario is shown in figure S2 in the supplementary information.

We do not model changes in climate prior to 2018, instead using Worldclim baseline estimates described as representative of conditions from 1960 - 1990 (Hijmans et al. 2005).

#### 2.1.5 Temperature suitability

We estimate the components of the temperature suitability index from YF-specific sources of information on extrinsic incubation period, vector mortality and bite rate for *Aedes Aegypti*, the urban vector of YF (Davis 1932; Tesla et al. 2018; Arran Hamlet et al. 2018; Mordecai et al. 2017). The extrinsic incubation period was estimated from the experimental results of Davis which were calculated specifically for YF in *Aedes Aegypti* (Davis 1932). We included bite rate data from both Mordecai et al. (2017) and Martens (1998) which both describe *Aedes Aegypti*. Finally, vector mortality was estimated from the experimental data of Tesla et al. (2018). Where data was provided in figure form, plots were digitised to extract the information. All data used for fitting the temperature suitability model are made available as Supplementary Information.

### 2.2 Models

We reformulate an established model of YF occurrence to accommodate nonlinear dependence on temperature and rainfall. We couple this with established results from a transmission model of serological status to estimate transmission intensity across the African endemic region at baseline/ current environmental conditions (Garske et al. 2014; Jean et al. 2018; Gaythorpe et al. 2019). Then, we project transmission intensity for four climate scenarios given projected changes in temperature and rainfall.

#### 2.2.1 YF occurrence

The generalised linear model (GLM) of YF occurrence provides the probability of a YF report at first administrative unit level for the African endemic region dependent on key climate variables. In order to assess the effect of climate change on YF transmission, we use the same methodology as (Garske et al. 2014; Jean et al. 2018; Gaythorpe et al. 2019); and incorporate covariates indicative of climate change that also have projections available in years 2050 and 2070 for different emission scenarios. As such, we omit enhanced vegetation index and land cover from the best fitting model of Garske et al. (2014) in favour of the temperature suitability index which depends on the average temperature, the temperature range and average rainfall. Temperature and rainfall are known to have implications on both the vectors of YF and the distribution of the non-human primate reservoir (Reinhold, Lazzari, and Lahondère 2018; Cowlishaw and Hacker 1997). However, the effect of temperature, particularly on vectors, is highly non-linear with increased mortality seen at very low and high temperatures; as such, we include the range in temperature as a covariate of our occurrence model as well as the non-linear temperature suitability index (Mordecai et al. 2017; Tesla et al. 2018). A full listing of covariates used in given in Supplementary table S2.

#### 2.2.2 Temperature suitability

We model suitability of the environment for YF transmission through temperature dependence. It has been shown that the characteristics of the virus and vector change with temperature (Brady et al. 2014; Kraemer et al. 2015; Mordecai et al. 2017; Tjaden et al. 2018). We model this using a function of temperature for the mosquito biting rate, the extrinsic incubation period and mortality rate for the mosquito which we combine to calculate the temperature suitability based on the Ross-MacDonald formula for the basic reproduction number of a mosquito-borne disease (Macdonald and others 1957). In the below, we focus on *Aedes Aegypti*.

The functional form used to model temperature suitability varies in the literature. We continue to use a form which can be parameterised solely from data specific to YF (Arran Hamlet et al. 2018; Garske, Ferguson, and Ghani 2013). However, alternative formulations have been published in the context of other arboviral infections (Mordecai et al. 2017; Ryan 2019; Brady et al. 2014, 2013; Tjaden et al. 2018).

Each input of the temperature suitability, *z*(*T*), is modelled as a function of average temperature where the individual thermal response follow the forms of Mordecai et al. The temperature suitability equation is as follows:

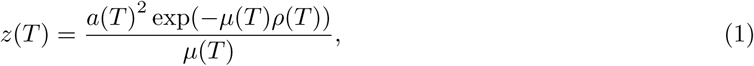

where *T* denotes mean temperature, *ρ* is the extrinsic incubation period, *a* is the bite rate and *µ* is the mosquito mortality rate. The thermal response models for *ρ, a* and *µ* follow Mordecai et al. (2017) as follows:

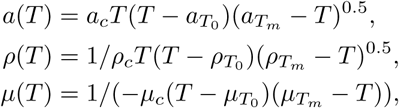

where the subscripts *T*_0_ and *T*_*m*_ indicate respectively the minimum and maximum values of each variable, and subscript *c* labels the positive rate constant for each model. The three resulting parameters for each model are estimated by fitting to available experimental data. The mortality rate *µ* is limited to be positive.

#### 2.2.3 Mapping probability of occurence to force of infection

We utilise previously estimated models of seroprevalence informed by serological survey data, demography and vaccination coverage information (Garske et al. 2014; Gaythorpe et al. 2019). The transmission intensity is assumed to be a static force of infection, akin to the assumption that most YF infections occur as a result of sylvatic spillover (Garske et al. 2014; Gaythorpe et al. 2019). The force of infection is assumed to be constant in each province over time and age. As such, we may model the serological status of the population in age group *u* as the following:

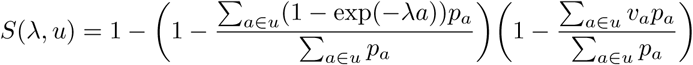

where *λ* is the force of infection, *p*_*a*_ the population in annual age group *a* and *v*_*a*_ the vaccination coverage in annual age group *a*. This provides us with estimates of force of infection in specific locations where serological surveys are available.

In order to estimate transmission intensity in areas where no serological survey data is available, we link the GLM predictions with seroprevalence estimates through a Poisson reporting process. The force of infection can be used to estimate the number of infections in any year. Thus, we may calculate the number of infections over the observation period. These will be reported with a certain probability to give the occurrence shown in the GLM. As such, we assume that the probability of at least one report in a province over the observation period, *q*_*i*_, depends on the number of infections in the following way:

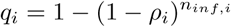

where *ρ*_*c*_ is the per-country reporting factor which we relate to the GLM in the following way:

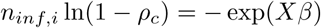

where *X* are the model covariates and *β*, the coefficients. The probability of detection can then be written in terms of the country factors, which are GLM covariates, *β*_*c*_, and *b*, the baseline surveillance quality calculated from the serological survey data:

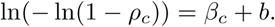

### 2.3 Estimation

We estimate the models of temperature suitability and YF report together within a Bayesian framework using Metropolis-Hastings Markov Chain Monte Carlo sampling with an adaptive proposal distribution (Andrieu and Thoms 2008; McKinley et al. 2014; Tennant, McKinley, and Recker 2019). The pseudo-likelihood contains components for the GLM of YF reports as well as the thermal response models and is given by the following:

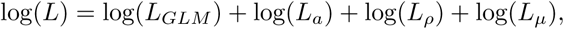

where log(*L*_*x*_) denotes the log likelihood of element *x*. The log likelihood for the GLM assumes that the binary YF occurrence data is Bernoulli distributed (Garske et al. 2014):

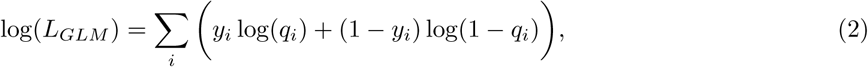

where *y*_*i*_ is the binary occurrence and *q*_*i*_ is the probability of at least one YF report in province *i*. We propogate uncertainty in the estimation of the GLM from the thermal response models as well as that from the seroprevalence into the resulting transmission intensity estimates.

The thermal response likelihoods are provided by an exponential distribution for bite rate, a Bernoulli distribution for mortality and a normal distribution for extrinsic incubation period.

The estimation, analysis and manuscript were all performed or written in R version 3.5.1, ridgeline plots were generated with packages ggplot2 and ggridges (R Core Team 2014; Wickham 2016; Wilke 2018; Garnier 2018).

### 2.4 Future projections

In order to assess future changes in force of infection, and thus disease burden, we incorporate ensemble climate projections of temperature change and precipitation. We assume that the force of infection is constant until 2018 and then changes linearly between 2018, 2050 and 2070, the years for which climate projections are available. Furthermore, in order to compare only the influence of changing population and force of infection, we assume that vaccination after 2019 is kept at the routine levels of 2018. As such, the results will not be affected by country specific preventive vaccination campaigns but, future burden will be over estimated as there are likely to be preventive and reactive campaigns in future. We estimate burden by calculating the proportion of infections who become severe cases and then, of those, the proportions that die, using published case fatality ratio estimates (Johansson, Vasconcelos, and Staples 2014). We compare burden estimates with a baseline scenario assuming the same demographic conditions and vaccination levels as the climate change scenarios but no change in climate variables (precipitation and temperature) over time.

## 3 Results

### 3.1 Model predictions for baseline scenario

Figure 2 (left) shows occurrence of YF across Africa from 1984 to 2018. Incidence is focused in the West of Africa and, more recently, Angola and the Democratic Republic of the Congo. The model predicts a high probability of YF report in these areas and reflects the general patterns of YF occurrence, see figure 2 for comparison. Model fit can be characterized by the Area Under the Curve (AUC) statistic (Huang and Ling 2005), which was 0.9004, similar to the original model formulation of Garske et al. (2014).

**Figure 2:**
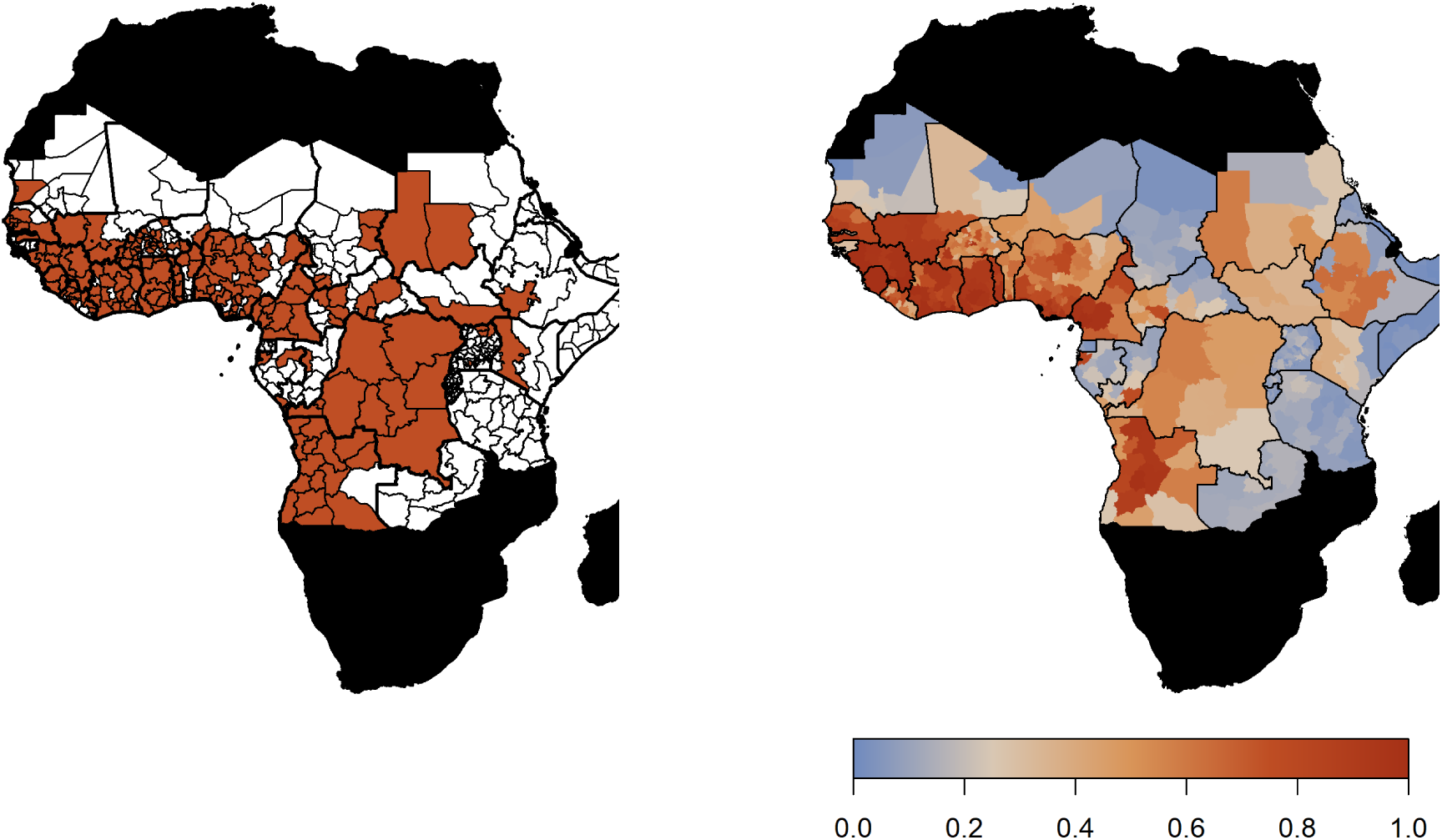
Observed YF occurrence (left) and median probability of a YF report predicted by the GLM.

The predicted probability of a YF report is positively informed by temperature suitability with the median posterior predicted distribution shown in figure 3a). This highlights the high suitability of countries such as Nigeria and South Sudan for YF transmission. In contrast, Rwanda, Burundi and areas of Mali and Mauritania have low average temperature suitability. The fit of the thermal response models is shown in the supplementary information, figures S11, S12 and S13.

**Figure 3:**
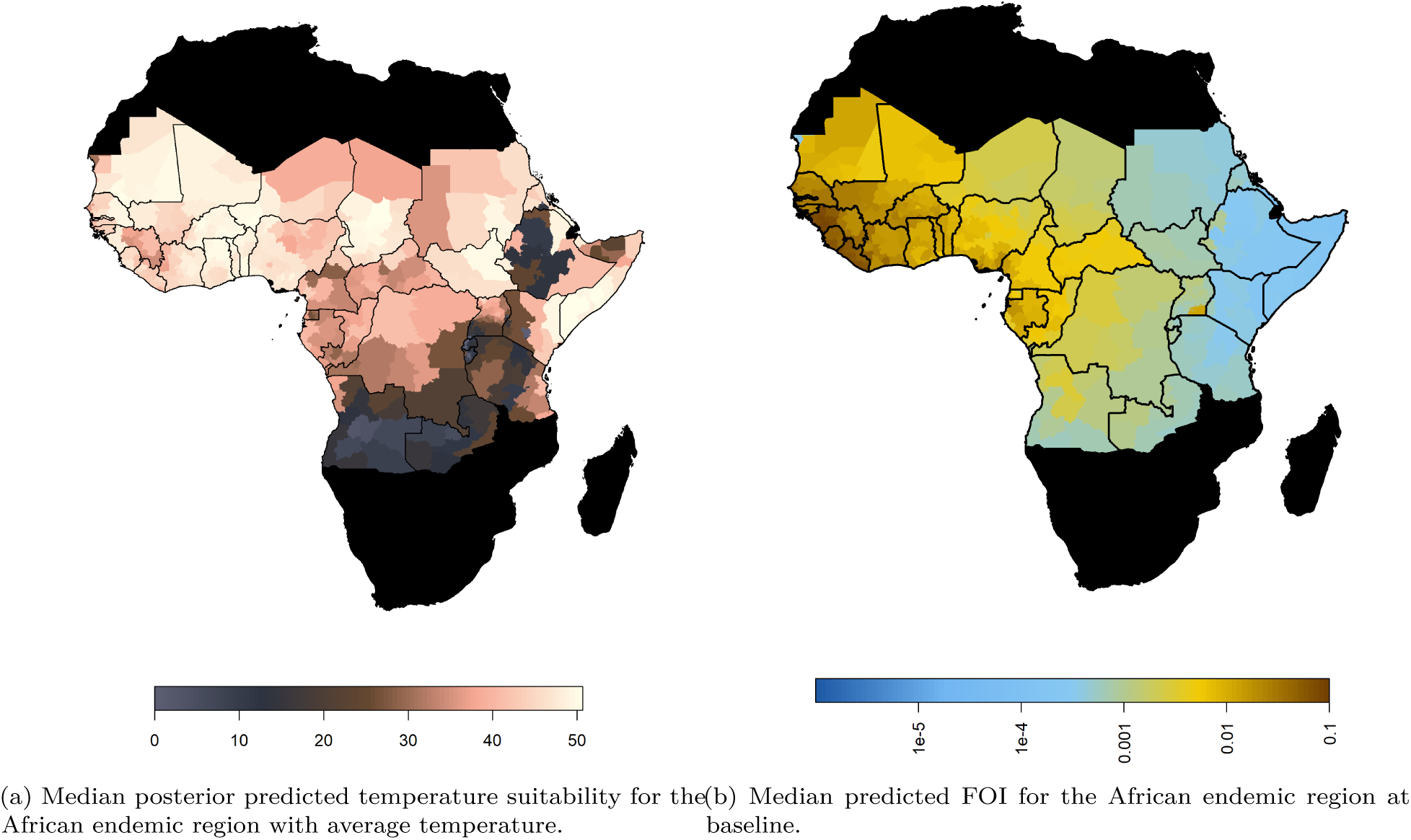
Median predicted model outputs for baseline scenario.

### Projected transmission intensity

Figure 3b) shows the median posterior predicted estimates of the force of infection for the baseline/ current scenario. When we incorporate the ensemble projections of temperature and precipitation change we see heterogeneous impacts on force of infection. Figure 4 shows the percentage change in median force of infection for the year 2070. Projections for 2050 are shown in figure S16 in the supplementary information.

**Figure 4:**
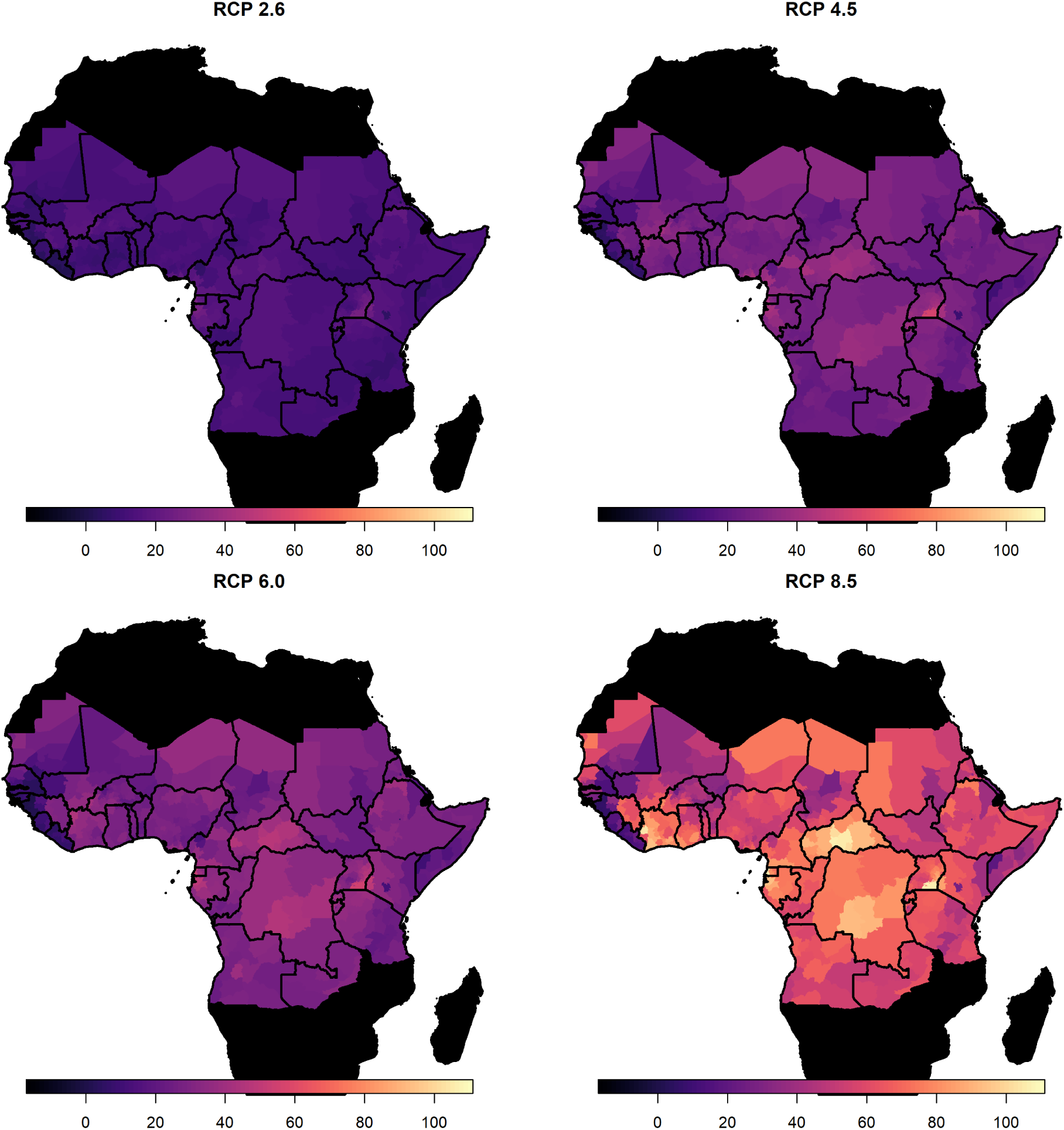
Percentage change in force of infection in 2070. Median predicted change in force of infection in the African endemic region in 2070 for the four emission scenarios.

The posterior distributions of predicted changes in force of infection in different African regions are shown in figure 5 (region definitions shown in figure S1). Projections for individual countries are given in supplementary information in figures S17 and S18. In West Africa, the predicted change is clustered around zero in the majority of scenarios; this is particularly the case for year 2050. However, due to wider uncertainty in 2070 and for RCP scenario 8.5 in general, there is a more discernible increase. In the East and Central regions, a predicted increase in force of infection is more apparent. Whilst the differences between 2050 and 2070 are difficult to see for RCP scenario 2.6, both peak above zero. In RCP scenarios 4.5, 6.0 and 8.5, the distinction between years is clear, particularly in 8.5, with the greatest increases seen in 2070 as temperatures are expected to continue to rise.

**Figure 5:**
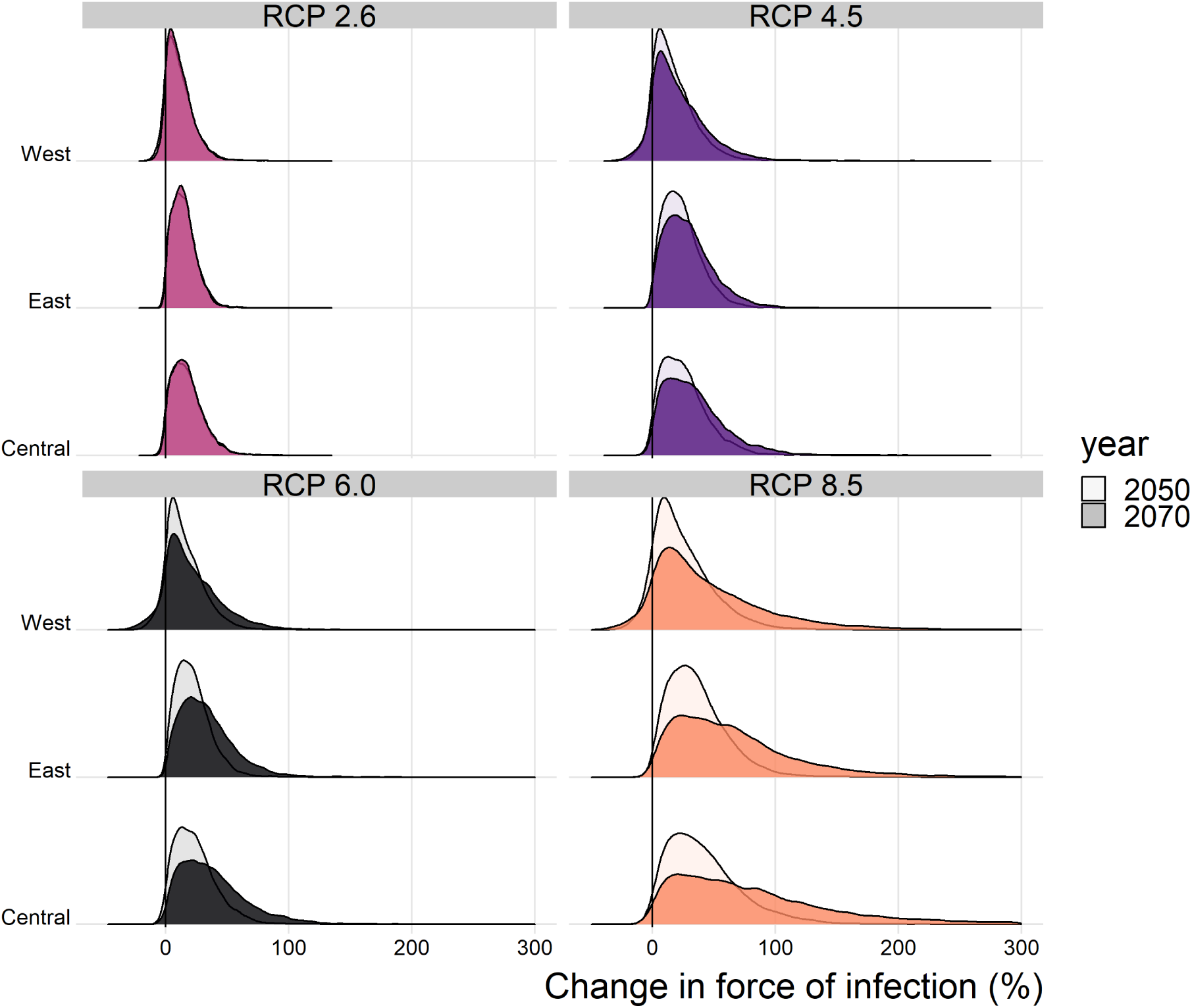
Predicted force of infection change (%) for each region of Africa, year and climate scenario.

When we examine the changes at country level, shown in the supplementary information, the changes are more heterogeneous. For RCP 2.6 Guinea Bissau the change in force of infection in 2070 is potentially broad, with a credible interval spaning zero: 10.3% (95%CrI [-33.2%, 96.3%]). Whereas, in Central African Republic, there is a notable increase by 87.1% (95%CrI [12.4%, 390.2%]).

### Projected burden

The projected percentage change in the annual number of deaths caused by YF across Africa is given in table 1; the projected annual deaths per capita for endemic countries are shown in figure 6 and in figure S19 in the supplementary information. These projections assume vaccination is static from 2019 onwards ie.e that only routine vaccination continues at 2018 levels. Aggregated numbers of deaths per country and region are shown in the supplementary information.

**Table 1:** Predicted percentage change in deaths in the African endemic region in 2050 and 2070 compared to the baseline/ current scenario.

| Year | Scenario | 95% CrI low | 50% CrI low | Median | 50% CrI high | 95% CrI high |
| --- | --- | --- | --- | --- | --- | --- |
| 2050 | RCP 2.6 | -2.36 | 4.49 | 10.84 | 18.58 | 37.91 |
| 2050 | RCP 4.5 | -2.40 | 7.32 | 16.71 | 28.16 | 57.43 |
| 2050 | RCP 6.0 | -2.78 | 6.79 | 15.49 | 25.86 | 51.85 |
| 2050 | RCP 8.5 | -2.17 | 11.03 | 24.92 | 41.84 | 88.33 |
| 2070 | RCP 2.6 | -0.74 | 4.11 | 9.99 | 17.03 | 34.10 |
| 2070 | RCP 4.5 | -2.76 | 7.77 | 19.28 | 33.56 | 71.08 |
| 2070 | RCP 6.0 | -4.56 | 8.63 | 21.35 | 36.70 | 77.70 |
| 2070 | RCP 8.5 | -2.90 | 16.08 | 39.57 | 72.43 | 178.63 |

**Figure 6:**
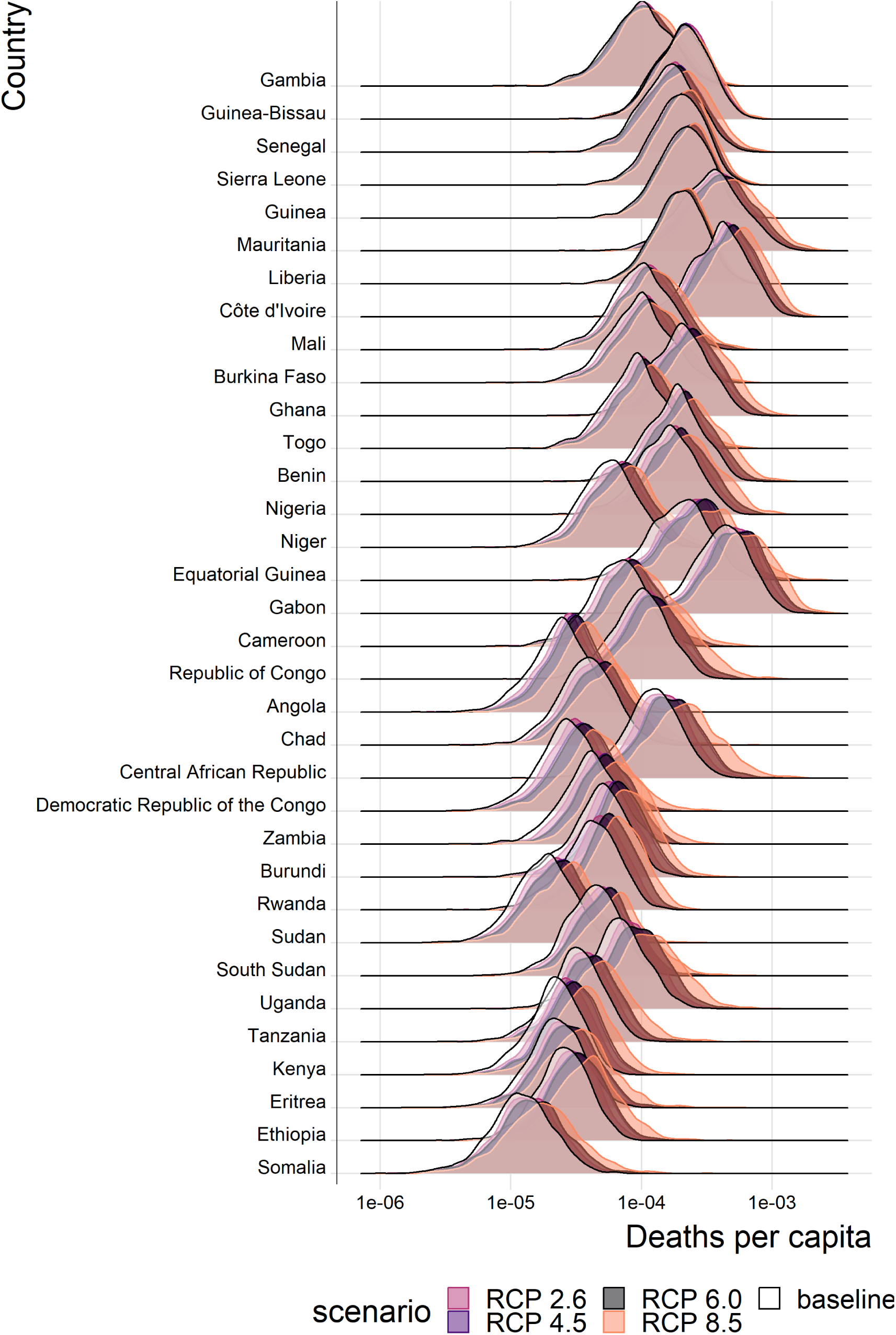
Posterior predicted annual YF deaths per capita for each country in the African endemic region in 2070. Countries are ordered by longitude.

While lower 95% credible intervals in table 1 are negative, the overall posterior probabilities that climate change will increase YF mortality are very high for each climate scenario. The probability that deaths will increase is 95.5% (95% CI [95.3%, 95.7%]) for RCP 2.6 in year 2070, rising to 95.9% (95% CI [95.7%, 96.1%]) for RCP 8.5 in year 2070, values for all scenarios and years are shown in supplementary table S4.

As with the force of infection projections, the most severe increases are seen for RCP scenario 8.5, especially in year 2070. The distinction between current projected deaths per capita and those under each RCP scenario are most clearly seen for countries in Central Africa, such as Central African Republic, and East Africa, such as Ethiopia. The four countries with the least distinct change, Liberia, Guinea, Sierra Leone and the Gambia, are all in West Africa, commonly thought to see the most intense YF transmission. As such, it appears that the most marked increases in burden are found in East and central Africa.

## 4 Discussion

We developed on an established model of YF occurrence and transmission to accommodate temperature and precipitation projections for four climate emissions scenarios. Non-linear dependence on temperature was incorporated by utilising a function of temperature suitability, informed by thermal response data for *Ae. Aegypti*. We jointly estimated parameters for the temperature suitability and occurrence models in a Bayesian framework, allowing us to quantify the uncertainty in our projections. We found, despite changes to the covariates used in the occurrence model compared with past work (Garske et al. 2014), that model fit remained good with a median AUC of 0.9004. This gave us some confidence in the suitability of the model for projecting the impact of climate change on YF transmission through to 2070, the last year for which climate emission scenario projections are available for temperature and precipitation.

The force of infection is projected to increase for the majority of countries in each scenario. Consistently, the Central African Republic is one of the countries most likely to see an increase in transmission, while Liberia and Guinea Bissau have much less clear projections. This highlights that the most severe proportional increases in force of infection are seen away from West Africa. However, as transmission is highest in West Africa, even a small relative increase of 3% (seen for Liberia in scenario RCP 2.6 in year 2050, see supplementary figures S17) could equate to a substantial increase in the projected absolute number of annual YF deaths.

In all scenarios there is a high probability that the number of deaths and deaths per capita will increase. The most marked changes are seen for RCP 8.5, the most severe emission scenario; however, changes are heterogeneous geographically with large proportional increases occurring in Central and East Africa. We expect the number of deaths per year to increase by 10.0% (95% CrI [-0.7, 34.1]) under RCP scenario 2.6 or 40.0% (95% CrI [-2.9, 178.6]) under RCP scenario 8.5 by 2070 (see table 1 for other values).

We assume that the force of infection changes linearly between 2018 and 2050, and between 2050 and 2070. The online supplementary video illustrates this by showing posterior saples of the change in deaths by region for all years between 2018 and 2070. For RCP scenario 2.6, deaths largely cease increasing after year 2050, in line with the assumption that RCP 2.6 represents the situation where contributing carbon activities peak by 2030; however, this scenario has been suggested to be ‘unfeasible’ (Mora et al. 2013; Vliet, Elzen, and Vuuren 2009). In RCP scenario 8.5, carbon contribution activities are assumed to continue increasing throughout the century and the consequent effects are seen in the projected number of deaths in East and Central Africa, with an acceleration after 2050.

Climate change will effect not only the magnitude of YF disease burden but also its distribution. This may lead to changing priorities with respect to vaccination. However, it is unclear whether the comparatively low proportional increase in burden seen for West Africa is due to more intensive vaccination or due to the limited increase in force of infection. Despite the ambiguity in mechanism, our results suggest that there could be drastic proportional increases in burden in East and Central Africa that may lead to greater vaccine demand in areas which have previously been of lower risk. Thus, whilst the countries experiencing the highest numbers of deaths will remain high risk, see figures S22 and S23 for the median distribution of deaths per year, countries such as Ethiopia and Somalia may become higher priority targets for vaccination.

Our analysis has a number of limitations. In order to utilise emission scenario projections we were limited to covariates with projections in 2050 and 2070, namely temperature and precipitation. This meant that we adapted our previous best-fit model (Garske et al. 2014) to include temperature range, temperature suitability and precipitation rather than enhanced vegetation and landcover. This change slightly reduced fit quality, giving an AUC of 0.9004 as opposed to to 0.9157 (Gaythorpe et al. 2019). Vegetation is a key factor determining habitat of non-human primates, an element that may not be captured by the temperature suitability index which focuses on the vector *Ae. Aegypti*. This omission may lead to an overestimation of the future burden as elements such as desertification and the impact of increasing frequencies of forest fires are not considered (Overpeck, Rind, and Goldberg 1990; Huang et al. 2016; James, Washington, and Rowell 2013).

Similarly, whilst the RCP scenarios model socio-economic and land-use changes, we do not explicitly include these aspects here (Van Vuuren et al. 2011). As such, we omit the human choices that may affect population distributions and behaviour, for example urbanization which has been shown to both reduce disease burden (Wood et al. 2017) and increase emergence of arboviruses (Gubler 2011; Hotez 2017). In the same way, while our model accounts for migration through use of the UN WPP population data, climate scenario-specific migration is not included in the model.

Data availability constrains aspects of our modelling approach. We use *Ae. Aegypti* and YF-specific datasets to inform the thermal response relationships and thus, temperature suitability index. However, some data, such as information on the extrinsic incubation period are severely limited; we use a dataset of experimental results from 1930s (Davis 1932). These data may be outdated due to current mosquito species potentially adapting to different climates as well as improved experimental procedures. This is a key data gap for YF and new experimental results concerning the extrinsic incubation period could provide valuable insight into the dynamics of the virus in mosquitoes today.

As further experimental data on thermal responses for *Ae. Aegypti* and other vectors of YF become available, the temperature suitability index developed here will be able to be enhanced. YF is known to have multiple vectors, each contributing to transmission cycles differently (Monath and Vasconcelos 2015), which are likely to have different thermal responses.

We focus only on a constant force of infection model which is similar to assuming the majority of transmission occurs as a result of zoonotic spillover. This assumption is supported by recent studies (Gaythorpe et al. 2019); however, the urban transmission cycle, driven by *Ae. Aegypti* plays a crucial role in YF risk and was responsible for recent severe outbreaks such as that in Angola in 2016. Incorporating climate projections into models that examine multiple transmission routes and thermal responses for multiple vectors, would produce a more realistic picture of how the dynamics of this disease may change with climate.

Climate change is projected to have major global impacts on disease distribution and burden (Mordecai et al. 2017; Huber et al. 2018; Kraemer et al. 2015). Here we examined the specific effects on YF and find that disease burden and deaths are likely to increase heterogeneously across Africa. This emphasises the need to implement and prepare for new vaccination activities, and consolidate existing control strategies in order to mitigate the rising risk from YF. Intervention through vaccination is the gold standard for YF, and new approaches are being implemented with respect to fractional dosing which is a useful resort to respond to urban outbreaks in case of vaccine shortage (Vannice, Wilder-Smith, and Hombach 2018). Yet, vaccination is not the only potentially effective control for YF, with novel vector control measures such as the use of Wolbachia showing promise and perspectives to improve clinical management or urban resilience (Rocha et al. 2019; World Health Organization 2017). Finally, in order to monitor and respond to changing transmission patterns, effective and sensitive surveillance will be essential.

## Data Availability

Data Availability: Public repository data: Vaccination coverage: coverage is available to download from the PoLiCi shiny app : https://shiny.dide.imperial.ac.uk/polici/. Serology surveys: There are seven published surveys used, available at DOI:10.1016/0147-9571(90)90521-T, DOI:10.1093/trstmh/tru086, DOI: 10.1186/s12889-018-5726-9, DOI: 10.4269/ajtmh.2006.74.1078, PMID: 3501739, PMID: 4004378, PMID: 3731366 Demographic data: Population level data was obtained from UN WPP https://population.un.org/wpp/, this was disaggregated using Landscan 2014 data https://landscan.ornl.gov/landscan-data-availability. Environmental data: This was obtained from LP DAAC: https://lpdaac.usgs.gov/ and worldclim http://www.worldclim.org/ Yellow fever outbreaks: These were compiled from the WHO weekly epidemiologic record and disease outbreak news https://www.who.int/wer/en/ and https://www.who.int/csr/don/en/. Data elsewhere: The data from the WHO YF surveillance database and from recent serological surveys from WHO member states in Africa underlying the results presented in the study are available from World Health Organization (contact: William Perea, or Laurence Cibrelus, or Jennifer Horton).

## Notes

### Competing Interest Statement

The authors have declared no competing interest.

### Funding Statement

Bill and Melinda Gates Foundation (Bill & Melinda Gates Foundation) Valid : Katy A. M. Gaythorpe, Tini Garske OPP1117543 Bill and Melinda Gates Foundation (Bill & Melinda Gates Foundation) Valid : Katy A. M. Gaythorpe, Tini Garske OPP1157270 Medical Research Council (MRC) Valid : Katy A. M. Gaythorpe, Arran Hamlet, Tini Garske, Neil M Ferguson MR/R015600/1 This work was carried out as part of the Vaccine Impact Modelling Consortium (www.vaccineimpact.org), but the views expressed are those of the authors and not necessarily those of the Consortium or its funders. The funders were given the opportunity to review this paper prior to publication, but the final decision on the content of the publication was taken by the authors. We acknowledge joint Centre funding from the UK Medical Research Council and Department for International Development. The funders had no role in study design, data collection and interpretation, or the decision to submit the work for publication.

